# Curtailing Covid-19 on a Dollar-a-Day in Malawi: Implications for the Ongoing Pandemic

**DOI:** 10.1101/2021.03.16.21253632

**Authors:** Iliana V. Kohler, Fabrice Kämpfen, Alberto Ciancio, James Mwera, Victor Mwapasa, Hans-Peter Kohler

## Abstract

Utilizing population-based data from the Covid-19 phone survey (N = 2, 262) of the Malawi Longitudinal Study of Families and Health (MLSFH) collected during June 2nd–August 17th, 2020, we investigate behavioral, economic and social responses to Covid-19 and focus on the crucial role that community leadership and trust in institutions play towards shaping these responses. We argue that the effective response of Malawi to limit the spread of the virus was facilitated by the engagement of local leaders to mobilize communities to adapt and adhere to Covid-19 prevention strategies. Village heads (VHs) played pivotal role in shaping individual’s knowledge about the pandemic and the adaption of preventive health behaviors and were crucial for mitigating the negative economic and health consequences of the pandemic. We further show that trust in institutions is of particular importance in shaping individuals’ behavior during the pandemic, and these findings highlight the pivotal role of community leadership in fostering better compliance and adoption of public health measures essential to contain the virus. Overall, our findings point to distinctive patterns of pandemic response in a low-income sub-Saharan African rural population that emphasized local leadership as mediators of public health messages and policies. These lessons from the first pandemic wave remain relevant as in many low-income countries behavioral responses to Covid-19 will remain the primary prevention strategy for a foreseeable future.

## 1 Introduction

Imagine the confluence of a novel pandemic with devastating global reach and consequences, a presidential election in a heated political climate after an annulment of the prior outcome, a fragile health care infrastructure, a long-standing HIV/AIDS epidemic and high prevalence of neglected tropical diseases and malaria (NTDMs), widespread poverty accompanied by crowded multi-generational living arrangements, and an economy structured around manual labor and in-person interactions that severely limit social distancing. Based on the experience in 2020, most would have expected Covid-19 to be unrelenting in this context: the virus spreading widely, and resulting in severe Covid-19-related morbidity and mortality as limited access to even the most basic treatments would prevent effective care for a large number of infected people. Not surprisingly, dire predictions about the effect of the pandemic on the “global poor” were abundant in the early phase of the pandemic (Gates 2020; Shuchman 2020; Van Zandvoort *et al*. 2020; Walker *et al*. 2020).

Yet, the pandemic unfolded differently in Malawi, a country ranked 174 out of 189 on the human development index (HDI) (UNDP 2000): Malawi is currently (January 19, 2021) ranked 186 in terms of reported cumulative Covid-19 cases per 1 Mio population (Worldometers 2020). While there is likely under-reporting of Covid-19 cases and deaths, the basic conclusion of a generally low Covid-19 incidence is corroborated by evidence of limited Covid-19 excess mortality (Bamgboye *et al*. 2020) or prevalence of Covid-19 symptoms (see below). In light of similar patterns in other low-income Sub-Saharan African countries (LIC SSA), and SSA countries currently reporting only 2.4% of all global Covid-19 cases and 2.7% of all deaths (WHO 2020), the possibility of a “sub-Saharan African Covid-19 puzzle” has been raised in both scientific journals and the popular media (Bearak and Paquette 2020; Maeda and Nkengasong 2021; Mbow *et al*. 2020; Mukherjee 2021). Explanations for these patterns, however, have been incomplete (Maeda and Nkengasong 2021). Demography, and specifically young population age structures, fewer chronic comorbidities linked to Covid-19 mortality and more circulating seasonal coronaviruses do not fully account for the low Covid-19 prevalence (Walker *et al*. 2020). Government spending on prevention and testing are also an unlikely explanation: in Malawi an estimated $213M required for effective pandemic responses faced $19M available for program implementation (The Republic of Malawi 2020), and vaccine roll-out is a still in its infancy with the government having committed to the AstraZeneca vaccine on February 1, 2021 (roll-out planned for March 2021).

In contrast, lessons learned from fighting the HIV/AIDS epidemic, Ebola, SARS and other community-spread diseases potentially facilitated swift and effective Covid-19 responses (Hargreaves *et al*. 2020; Paintsil *et al*. 2020). Consistent with this hypothesis, there is a negative relationship in SSA between prior death rates for neglected tropical diseases and malaria (NTDMs) and the estimated cumulative incidence of Covid-19 per 100k population (Figure 1). Motivated by this aggregate pattern, we argue in this paper that the effective response of Malawi to limit the direct health impacts of the Covid-19 pandemic was facilitated by the prior experience in fighting epidemics and community-spread diseases, and particularly by the combination of two key factors: (1) an early recognition by the government and individuals, including in rural areas, of the severity of the pandemic and its potentially dire health consequences, and (2) an effective response and engagement of local leaders to mobilize communities to adapt and adhere to Covid-19 prevention strategies.

**Figure 1.**
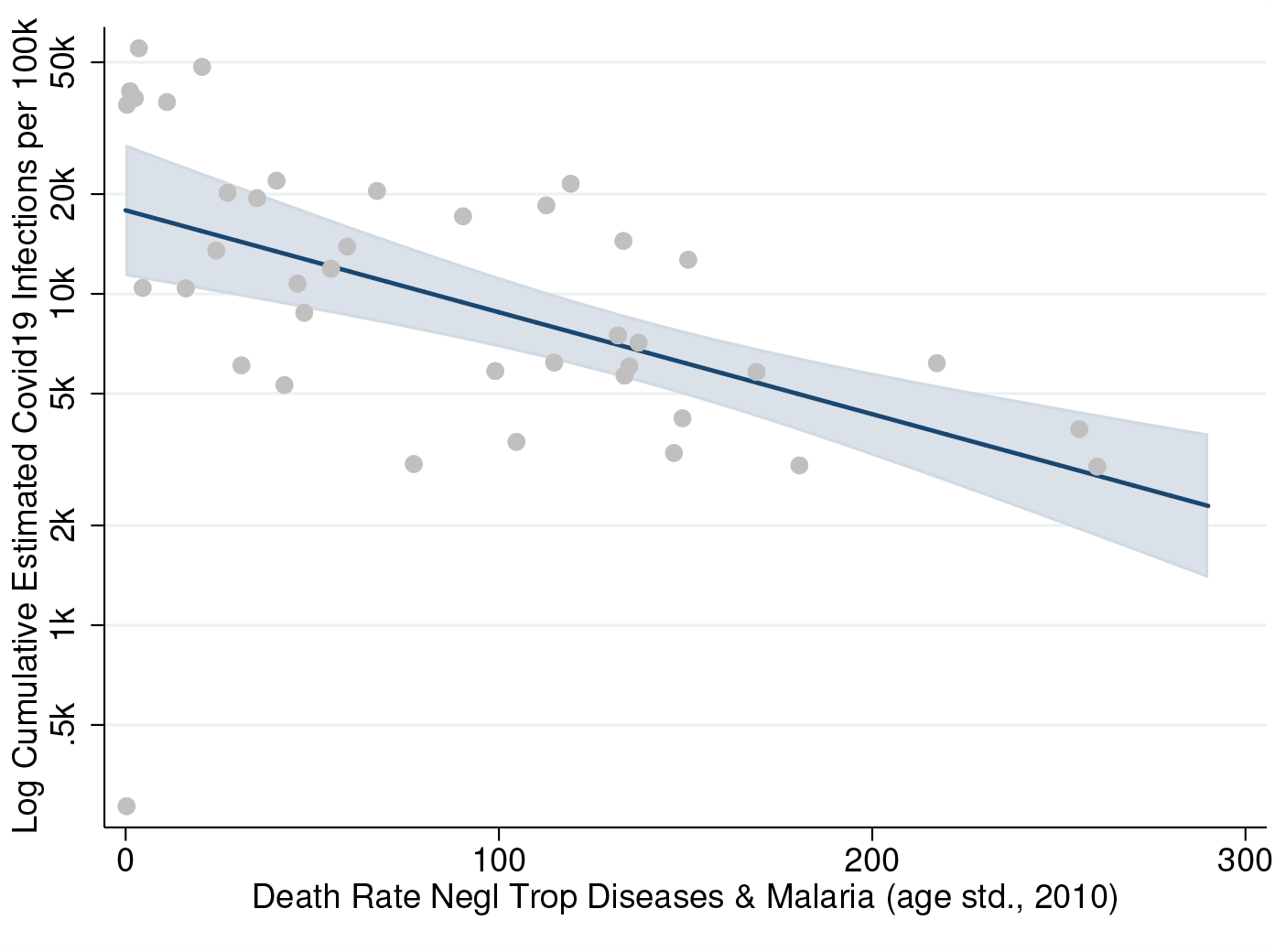
Cumulative Covid-19 incidence and prior mortality due to neglected tropical diseases and malaria (NTDM) *Notes:* Marginplot of the relationship between estimated cumulative Covid-19 incidence (data as of March 6, 2021), provided by IHME (IHME 2021)) and age-standardized death rate from neglected tropical diseases and malaria (NTDM) in sub-Saharan African countries (based on GBD 2019 (GBD 2019 Diseases and Injuries Collaborators 2020)). IMHE provides Covid-19 estimates for 40 SSA countries. For estimating the regression line, country observations are weighted by population size. Estimated cumulative Covid-19 incidence covers the period until 28 Feb 2021. The relationship is robust and holds for different years (2000 or 2010) and with controls for age structure and overall mortality level (all-cause age-standardized death rate); see Supplemental Table S1). A modest negative association is also found for mortality due to enteric infections, while prior HIV and STI mortality is positive correlated with Covid19 outcomes (Supplemental Table S1). No association is found with prior mortality due to respiratory infections and all infectious diseses overall (Supplemental Table S1).

Our study also affirms that meaningful involvement of local communities and local leadership can shape the population-wide response to Covid-19 and mitigate the secondary consequences of the pandemic. Specifically, supporting local leadership engaged in building social capital (i.e., disease knowledge, adoption of preventive health behavior), trust in governmental and health authorities and their health policies, can amplify public health messages (Chan 2014; Hargreaves *et al*. 2020; Hopman *et al*. 2020; Kao *et al*. 2021; Nuwagira and Muzoora 2020) and foster appropriate individual behavioral, social and economic responses to the Covid-19 pandemic, and potentially alleviate its impacts on communities such the ones we study in rural Malawi.

Our data collected between June 2nd–August 26, 2020, cover a critical period during which Covid-19 peaked and began to decline, shaping the long-term trajectory of the pandemic in Malawi. During this period, traditional community leaders (village heads, VHs) have been able to serve as critical interlocutor between the governmental policies and information dissemination on one side, and individuals’ and communities’ responses on the other side. Leveraging accumulated experience from combating prior epidemics, community leadership structures were critical to shaping and sustaining the population-wide response to Covid-19. With expected long delays until vaccinations will be administrated to the majority of the population in LIC SSA, and ongoing risks of pandemic relapses due to more easily spreading Sars-Cov-2 variants, the factors contributing to Malawi’s successful early response provide a template for the required prolonged effort to contain the pandemic in low-income countries through behavioral change and community mobilization.

## 2 Background

Malawi initiated a focused Covid-19 information campaign in February 2020, about two months before the first case was reported in the country. A special cabinet committee on coronavirus was established in March, and a state of emergency was declared shortly thereafter, resulting in the implementation of restrictive social distancing measures (including school closures at all levels, ban on public events and gatherings limited to 100 people, reduction in public transport capacity and border closures). A 21-day national lock-down beginning April 18th, however, was not implemented as a court injunction deemed it unconstitutional and potentially causing a severe nutritional crisis. The subsequent phase of the pandemic coincided with a volatile period of political uncertainty, polarization and a heated election campaign, ultimately leading to a new government on June 28th (BBC News 2020; Chirwa *et al*. 2020). Yet, irrespective of which administration was in place, the government maintained a focus of its Covid-19 campaigns on risk communication and community engagement following a similar approach employed to address the HIV/AIDS epidemic. Public health messages were disseminated via national radio, interactive phone text messages through both phone network suppliers, distribution of printed materials, community awareness meetings, and others. During April–May, mobile van units for the distribution of Covid-19 information and educational materials were mainly employed in the urban areas, expanding nationally only after the election and intensifying in July as cases rose. Importantly, the government transition in June significantly increased how factually truthful individuals perceived the government to be about Covid-19 (Figure 2).

**Figure 2.**
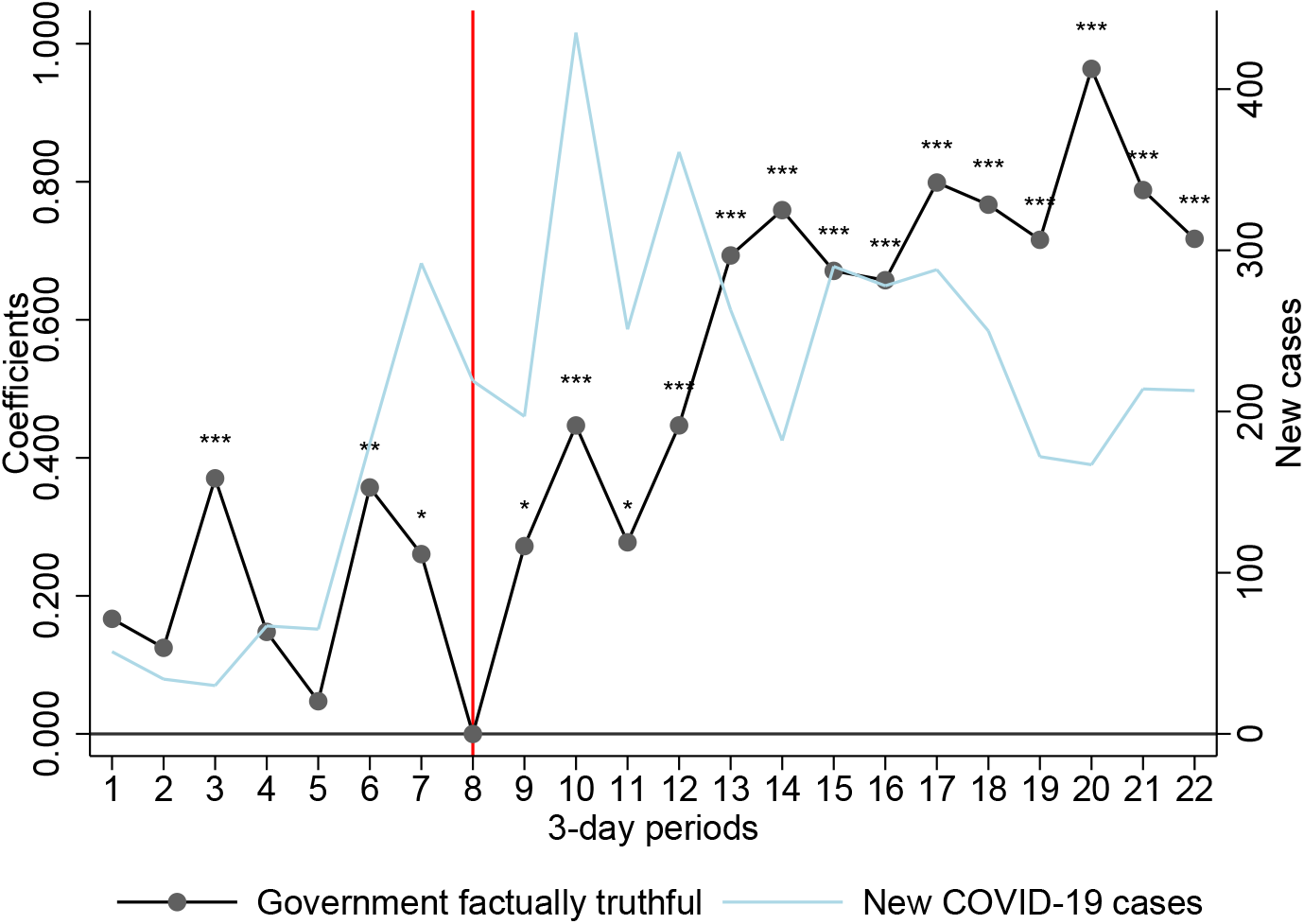
Pre- and post-election trust in the government’s messaging about Covid-19. *Notes:* Red line denotes change in government. Black line shows the coefficients associated with time dummy variables (3-day period time) estimated from a regression of survey responses on the government’s truthfulness on a set of control variables (age, gender, schooling, region, ^*^ *p* < 0.1, ^* *^ *p* < 0.05, ^* * *^ *p* < 0.01). Superimposed is the Covid-19 incidence data reported by the Ministry of Health (Public Health Institute of Malawi 2020). Trust in the government was assessed with the question “*How factually truthful do you think your country’s government has been about the Covid-19 epidemic?* “with responses measured on a Likert scale ranging from 1=‘very untruthful’ to 5=‘very truthful’.

It is important to acknowledge that, despite the relatively low prevalence of Covid-19 infection and death rates, the pandemic has exacerbated economic and other health challenges in SSA LICs. Poverty levels in SSA quickly aggravated through the worldwide stall in the global economy. Food insecurity and malnutrition increased, and children’s immunizations decreased sharply to levels last seen in the 1990s (BMGF 2020; Egger *et al*. 2021). Through these secondary effects, the Covid-19 pandemic has the potential to result in devastating health and social crises even if Covid-19 infections were to remain relatively curtailed (A Madhi *et al*. 2020; BMGF 2020; Roberton *et al*. 2020). A striking dichotomy therefore characterizes Covid-19’s impact on in Malawi and other SSA LICs: economic and related social and secondary health impacts have been severe (Buonsenso *et al*. 2020; Burger *et al*. 2020; Egger *et al*. 2021; Kanu 2020; Roberton *et al*. 2020), while the direct health consequences of the Covid-19 pandemic—Covid-19 infections and related mortality—have been substantially more modest than many observers expected at the beginning of the pandemic (Musa *et al*. 2021).

## 3 Sample Characteristics and Methods

<We use data from a Covid-19 Phone Survey that was conducted as part of the Malawi Longitudinal Study of Families and Health (MLSFH) (Kohler *et al*. 2015) during June 2nd—August 26th, 2020. The target sample included MLSFH respondents, village heads (VHs) and health care providers (HCPs) serving the three MLSFH study areas who were interviewed during 2017–2019 and for whom at least one phone number was available. Out of 3,172 eligible MLSFH respondents (excluding HCPs), 2,262 were successfully interviewed (71% response rate), with the response rate being higher among younger study participants (71% to 78% for ages 25–64 years), while only 62% among those age 65-74 years and 47% of those age 75+ years were successfully surveyed. Survey participants were on average 49 years old and about 42% were men. The majority of the respondents were currently married (84.7%), and had finished standard/primary level of schooling (67.8%). About equal proportions of respondents lived in the central (Mchinji) and northern (Rumphi) study districts (35-36%), 25% were located in the southern (Balaka) district, and about 4% were residing in other parts of Malawi as a result from migration out of the three MLSFH study areas (Supplemental Table S2). The survey collected a comprehensive range of information on Covid-19 related topics ranging from knowledge about transmission pathways and behavioral responses to reduce infections, experience of current and past Covid-19 infection symptoms among respondents and members of their households, impact of the pandemic on economic and social well-being, trust in institutions, village-level responses to the pandemic, to subjective well-being and mental health during the pandemic. Our analyses are primarily based on regression models (linear or ordered probit) that control for gender, age, schooling, region of residence and time (survey day fixed effects to control for systematic difference across days).

Consistent with the narrative of a relatively successful containment of the Covid-19 pandemic in Malawi and other SSA LICs, very few MLSFH respondents reported the experience of any Covid-19-related symptoms at the time of the interview (3.7% had fever, 14.4% had dry cough, 1.9% had shortness of breath, and only 0.3% reported having all these three symptoms simultaneously; very few reported symptoms among household members; Supplemental Table S2). Only four respondents reported having been diagnosed with Covid-19, two of whom based on a coronavirus test. Similarly low prevalence of Covid-19 symptoms in mid-2020 and relatively modest excess mortality due to Covid-19 have also been documented in other rural SSA LICs’ communities (Bamgboye *et al*. 2020; Banda *et al*. 2020).

Despite low prevalence, however, the impact of Covid-19 on daily lives and livelihoods was apparent even at this early stage of the pandemic. About 77.1% of respondents had reduced their non-food expenditures (e.g., expenses related to children’s schooling, agriculture, transportation and entertainment), 19.2% had reduced food consumption, 16.3% reduced health expenditures as a result of the pandemic. 55.3% reported an overall worsening economic situation, and 22.2% reported increased food-related worries during the last 12 months. The pandemic also exacerbated concerns about access to health care, including HIV treatment, children’s vaccinations, and treatment for non-communicable diseases (NCDs) (Supplemental Table S2).

Because of their eminent roles in all village-related matters, including monitoring compliance with public health measures and their authority to sanction dissent behavior (Kao *et al*. 2021), the MLSFH 2020 Covid-19 Phone Survey elicited information on the pandemic-related activities of respondents’ village heads (VHs). In this analysis, we define a VH as “*socially active*” if he/she had instructed village residents to implement social distancing measures (cancel village meetings, keep distance from other people during activities outside of the household (i.e., when fetching water), stop public works or recreational activities on common playgrounds). We define a VH as “*economically active*” if he/she had given instructions to create a village fund for emergency purposes or redistribute resources (i.e., food, money, medical supplies) to the most vulnerable residents. 87% of respondents reported VHs who were socially active only, 25% VHs who were economically active only, 24% VHs who were both socially and economically active, and 13% VHs who were neither socially nor economically active. The MLSFH longitudinal information showed that whether VHs are active or inactive during the Covid-19 pandemic is partially related to the extent to which these rural communities have experienced adversity in the past. For example, Supplemental Table S3 shows that respondents living in villages that experienced relatively high number of negative economic shocks between 2008 and 2010 were more likely to have a socially active VH more than a decade later.

## 4 Results: Curtailing the pandemic at a Dollar-a-day

### 4.1 Perceptions of disease risk: Covid-19 vs. HIV

A possibly important contributor to Malawi’s low Covid-19 incidence was the widespread perception of Covid-19 as a severe health threat early in the pandemic when local incidence was still very low (Figure 3). To put Covid-19 risk perceptions into perspective, we compare them to analogous perceptions for the same respondents measured around the peak of the HIV epidemic (2006) when antiretroviral treatment (ART) was not yet available in the MLSFH study areas. Respondents (correctly) estimated that the 2006 prevalence of HIV in their communities was substantially higher than the Covid-19 prevalence in mid-2020 (Panel A in Figure 3). At the same time, they perceived a much higher likelihood to be infected with Covid-19 as compared to HIV, and they perceived a rapidly increasing risk of Covid-infection as the pandemic unfolded during mid-2020 (Panel B).

**Figure 3.**
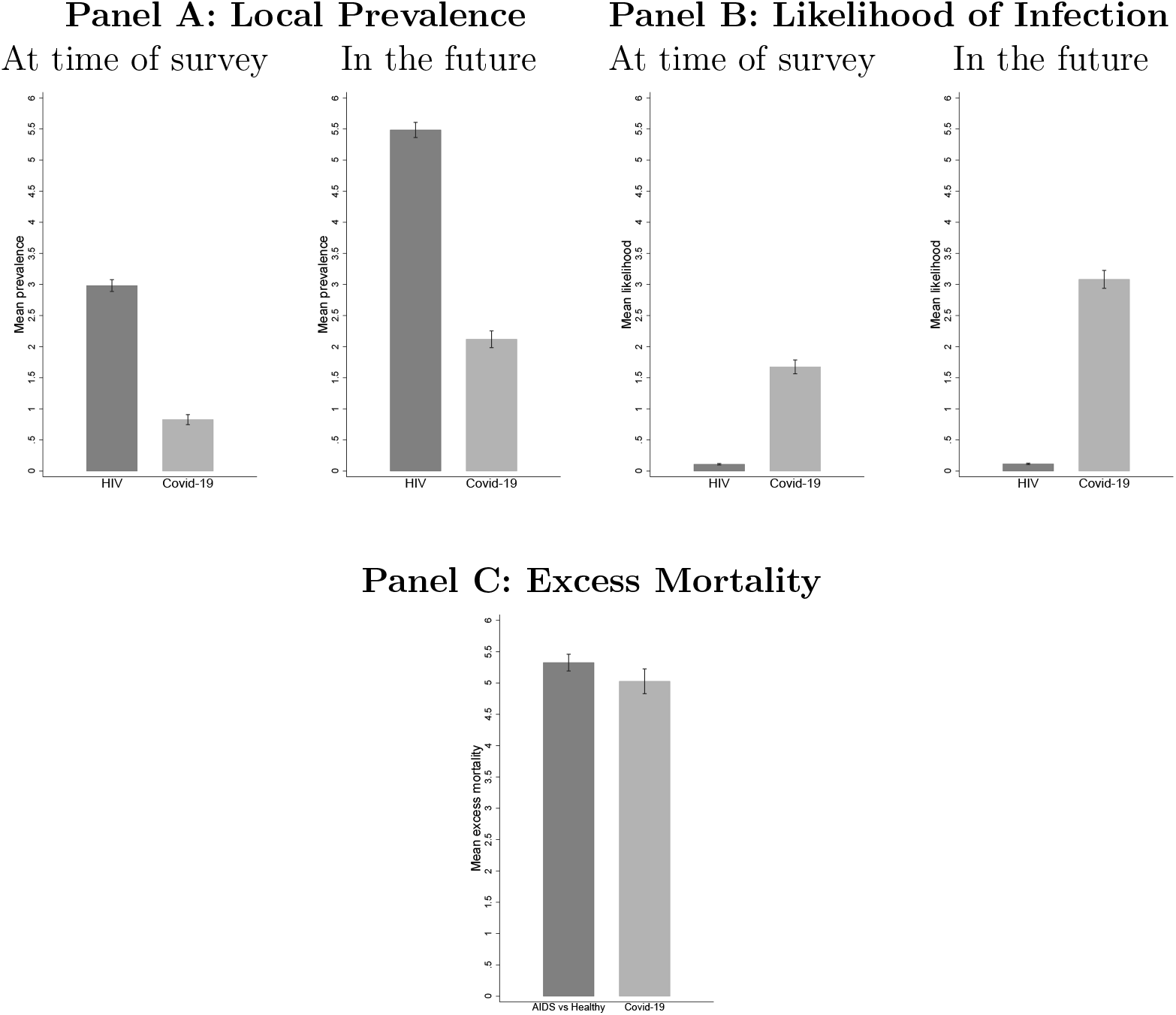
Comparison of Covid-19 (mid-2020) and HIV (2006) Perceptions. *Notes:* **Panel A**: Perceptions of local prevalence are estimated based on the question “*If we took a group of 10 people from this area—just normal people who you found working in the fields or in homes—how many of them do you think would now have coronavirus (Covid-19)?* “(2020), and the analogous question that was asked about persons infected with HIV in 2006. “In the future” refers to 3 months for Covid-19 and 5 years for HIV in 2006. **Panel B**: Likelihood if infection for Covid-19 (2020) and HIV (2006) is obtained using a method to elicit subjective probabilistic expectations that has been implemented in the MLSFH since 2006 (Delavande and Kohler 2009). In the 2020 phone survey, this question was worded as “*Out of 10, tell me the number of peanuts that reflects how likely you think it is that you are infected with coronavirus (Covid-19) now?*,” where each peanut represents a 10% chance. An analogous question was asked about the likelihood of being infected with HIV in 2006. “In the future” refers to 3 months for Covid-19 and 2 years for HIV in 2006. **Panel C**: Perceived excess mortality for HIV was estimated using the difference between the probability of a hypothetical person dying in 5 years if sick with HIV/AIDS minus the probability of a health person dying in 5 years. Perceived excess mortality for Covid-19 was estimated as the probability of dying *of Covid-19* conditional on being infected.

Combining data on perceived prevalence and perceived mortality conditional on being infected with Covid-19 or HIV allows an approximation of respondents’ *excess mortality risk* as a result of Covid-19 in 2020 and HIV in 2006 (Panel C in Figure 3). Importantly, Panel C suggests that perceived excess mortality in mid-2020 due to Covid-19 was of similar magnitude as that of HIV near the peak of the epidemic in 2006 before the country-wide introduction of ART. This finding indicates that rural Malawians very early in the pandemic attributed a substantial health and mortality risks to Covid-19, facilitating the adaption of preventive measures and the development of community responses. These perceptions of high excess mortality due to Covid-19 in Malawi are in stark contrast to the relatively low perceived excess mortality among US residents early in the pandemic (Ciancio *et al*. 2020).

### 4.2 Role of local leadership for behavioral responses to Covid-19

Relatively good knowledge about Covid-19 was widespread already in the early phase of the pandemic. 34.2% of respondents were able to list the main infection symptoms (dry cough, fever and difficulties breathing), and 68% knew that infected people can be asymptomatic. Almost all (86.3% to 95.6%) knew the primary transmission pathways. Similarly, protective and social distancing measures were widely known and followed by the survey participants: 70% reported decreased time spent close to people outside of their household, 91% had avoided close contact to other people, and 72.6% reported staying at home to prevent infection (see Supplemental Table S2 for additional indicators). Evidence of high Covid-19-related knowledge and social distancing measures have already been documented in Malawi (Banda *et al*. 2020; Fitzpatrick *et al*. 2021). Variation in respondents’ knowledge is importantly related to the local leadership, and in particular village heads (VHs): respondents whose VHs were socially and/or economically active were more likely to own and/or wear face masks, had higher knowledge of Covid-19 symptoms, knew more risk-reducing behaviors for Covid-19 infection (i.e., measured by the RR score) and were taking more actions to prevent infections (i.e., as measured by the Action Score) (Table 1). These associations were particularly large for individuals whose VHs were socially active. Differences between socially and/or economically active VHs also emerge once Covid-19 prevention strategies are classified as “*low cost* “(e.g., washing hands, avoiding shaking hands, avoiding close contacts with people outside of the household) and “*high cost* “(staying at home, or decreasing time spent with people outside of the household, with the latter baring high economic costs and being difficult to implement in subsistence agricultural societies) (see Supplemental Table S4). Respondents were more likely to implement both low and high costs measures if they had a socially active VH, while economically active VH is only associated with adopting low costs measures.

**Table 1:** Associations between village head’s (VH) characteristics and behavioral responses to Covid-19.

|  | Wear<br>face masks<br>(1) | HH owns<br>face masks<br>(2) | Symptoms<br>score<br>(3) | RR<br>score<br>(4) | Action<br>score<br>(5) |
| --- | --- | --- | --- | --- | --- |
| VH socially active | 0.157***<br>(0.029) | 0.153***<br>(0.028) | 0.289***<br>(0.089) | 0.203***<br>(0.065) | 0.745***<br>(0.095) |
| VH economically active | 0.115***<br>(0.023) | 0.097***<br>(0.023) | 0.116<br>(0.073) | 0.031<br>(0.043) | 0.261***<br>(0.054) |
| Observations | 2131 | 2132 | 2125 | 2127 | 2128 |

### 4.3 Role of local leadership in mitigating the consequences of Covid-19

VHs’ activities were importantly related to how respondents tried to mitigate the health and economic consequences of the pandemic (see Supplemental Table S5). Respondents with economically active VHs were more likely to reduce non-food and health expenditures at this early stage of the pandemic, possibly as a result of VHs advising respondents to smooth their consumption and contribute to public funds to help other villagers experiencing economic difficulties (Column 1 in Table S5). There is no association between having an economically active VH and a decrease in food expenditures, while the association exists for socially active VHs (Column 2). This difference possibly reflects the fact that food expenditures are more inelastic and individuals who felt financially more stable thanks to the behavior of their VHs did not have to reduce their food consumption. This pattern is also supported by the negative association between economically active VHs and the increase in food worries over time (Column 5) and in the frequency of eating less food than needed (column 6) since 2019. These associations are not statistically significant for socially active VHs. Respondents whose VHs were socially or economically active were more likely to borrow money than others (Column 4), suggesting the ability of VHs to provide stable and less risky environment for individuals to make financial decisions during the pandemic. Having an economically active VHs was negatively associated with respondents’ worries about access to health care during the pandemic (Table S7), including HIV testing, pre/post-natal care, vaccinations, access to contraception, while socially active VHs were not systematically associated with mitigating health care related worries (see Supplemental Table S6).

### 4.4 Trust in institutions and well-being during Covid-19

Respondents expressed relatively high levels of trust and confidence in the two institutions mostly in charge for the country’s response to Covid-19: the health care system as represented by the health care workers and the government. About 80.2% of the respondents trusted that health care workers “do what it takes” to minimize the negative impact of the Covid-19 pandemic, and the majority of respondents (61.8%) thought of the Malawian government as being factually truthful about the pandemic, with an increasing trend after the elections (Figure 2). Perceiving the government as truthful is associated with lower worries about Covid-19 affecting access to health care (Supplemental Table S7). On average, only 21.1% of respondents perceived the government as having been untruthful with pandemic-related information, and respondents who rated the government as factually untruthful about Covid-19 were less likely to implement social distancing measures (Supplemental Table S8). Similarly, respondents who distrusted health care workers had lower probability to adopt social distancing measures or preventative health behaviors compared to those who consider health care workers neither truthful not untruthful with respect to the Covid-19 response (Supplemental Table S9). Finally, respondents who perceived the government as truthful or trusted the health care workers generally reported higher levels of subjective well-being and self-reported health, suggesting that they were subjectively enduring the pandemic better than others. Except for depression, these associations persisted when measuring changes between the most recent pre-pandemic measures of well-being or subjective health and the corresponding outcomes measured during 2020 (Supplemental Table S10).

Finally, our results in Table 2 emphasize the important role socially active VHs play for building trust in the government and the health care system (health care workers). Specifically, respondents who lived in villages with socially active VHs expressed higher trust in the health care workers to deal with the pandemic and also perceived the government being truthful with Covid-19 messages. In contrast, this association was not established for economically active VHs.

**Table 2:** Association between village head’s (VH) characteristics and trust towards institutions.

|  | Trust in<br>HW<br>(1) | Trust in<br>HW<br>(2) | Trust in<br>HW<br>(3) | Gvt<br>truthful<br>(4) | Gvt<br>truthful<br>(5) | Gvt<br>truthful<br>(6) |
| --- | --- | --- | --- | --- | --- | --- |
| VH socially active | 0.333***<br>(0.089) |  | 0.327***<br>(0.090) | 0.263***<br>(0.078) |  | 0.249***<br>(0.080) |
| VH economically active |  | 0.080<br>(0.074) | 0.030<br>(0.076) |  | 0.100<br>(0.064) | 0.065<br>(0.065) |
| Observations | 2130 | 2130 | 2130 | 2129 | 2129 | 2129 |

*Note:* Estimates are derived from ordered probit regressions with robust standard errors reported in parentheses (\* $p < 0.1$ , \*\* $p < 0.05$ , \*\*\* $p < 0.01$ ). All regressions control for sex, age (dichotomous variables for age 19-34, 35-44, 45-54, 55-64, 65-90) and education (dichotomous variables for "never attended school", "finished standard" and "finished form and above"), and include region and time (in days) fixed-effects. We define a VH as being “socially active” if he has instructed respondents to cancel village meetings, keep distance from other people while fetching water, stop public works or stopping recreational activities, such as soccer on the playground. We define a VH as being “economically active” if he has instructed respondents to creating a village fund for emergency purposes or redistribute resources (food, money, medical supplies) to the most vulnerable members of the village community. “HW” stands for health care workers. Outcome variables take three possible values: 0 (very untruthful or somewhat untruthful/strongly distrust or somewhat distrust), 1 (neither truthful nor untruthful/neither trust nor distrust) and 2 (very truthful or somewhat truthful/strongly trust or somewhat trust).

## 5 Summary and Discussion

How did Malawi, a poor country where more than half of the population lives on less than one dollar a day, succeed in curtailing the Covid-19 pandemic with relatively low rates of infections and excess mortality? While multiple mechanisms are likely to contribute to this pattern, including a favorable (young) population age structure, effective behavioral and institutional responses during the early phase of the pandemic almost certainly deserve substantial credit. This pattern is consistent with the observation that the timely and decisive handling of the Covid-19 pandemic and a joint continental strategy have been an important factor that may have influenced the pandemic’s trajectory across the African continent(Maeda and Nkengasong 2021; Rosenthal *et al*. 2020). Focusing on the period June–August 2020 that was critical for shaping the longer-term trajectory of the disease in Malawi, we document that this effective behavioral and institutional response to Covid-19 in the country is related to four factors. First, despite a presidential election and government transition in mid-2020, the country adopted early in the pandemic a sustained prevention-focused information campaign that emphasized community engagement and dissemination of risk-reduction strategies through multiple channels to ensure a broad reach of the primarily rural population. In mid-2020, our predominantly rural MLSFH respondents reported fairly high knowledge of Covid-19 disease symptoms, transmission pathways, appropriate behavioral responses, and they reported widespread compliance with social distancing and other preventive behaviors. These findings are consistent with previously documented patterns in the same context (Banda *et al*. 2020; Fitzpatrick *et al*. 2021).

Second, the widespread adaption of preventive behaviors was likely facilitated by the recognition of Covid-19 as a severe health risk that can entail significant mortality risks. Drawing on longitudinal MLSFH data from 2006–2020, comparisons of subjective mortality expectations indicate that study respondents associated an excess mortality risk with Covid-19 that is of similar magnitude of that resulting from HIV/AIDS near the peak of the HIV-epidemic when antiretroviral treatments were not yet widely available. These perceptions of high excess mortality due to Covid-19 in Malawi are in stark contrast to the relatively low perceived excess mortality among US residents early in the pandemic (Ciancio *et al*. 2020).

Third, our findings point to the crucial role of local leadership (village heads, VHs) to mitigate the impact of Covid-19 in poor rural subsistence communities. Specifically, we show that respondents who live in villages with socially active village heads are more knowledgeable about the pandemic and more likely to adopt preventive health behaviors. Importantly, individuals living in villages with economically active village heads were less likely to report food worries, suggesting a better ability to smooth their food consumption as a response to Covid-19. Variation in the extent to which village heads were active during the early-phase of the Covid-19 pandemic does not seem to be random. On the contrary, respondents who live in villages with relatively high number of negative economic shocks reported between 2008 and 2010 were more likely to have a socially active village head more than a decade later during the Covid-19 pandemic (Supplemental Table S3). This finding is of particular importance since it provides evidence that lessons learned from the past, not only from fighting the HIV/AIDS epidemic and other community-spread diseases but also from surviving economic hardship and instability during periods of community-wide adversities, have facilitated effective Covid-19 responses that contribute to reducing the negative consequences of the pandemic.

Fourth, our findings point to the important role of the community leaders (i.e., VHs) as being critical to build trust in national institutions such as the health care system and the government during the pandemic, which in turn can result in better compliance and adoption of public health measures to contain the virus. For instance, we showed that respondents who distrust the health care system and the government are less likely to follow social distancing practices, a behavior that is detrimental to efforts to contain the pandemic (Tables S8 and S9). This finding is important as broad public support, trust in institutions, and culturally informed and credible public health communication are often emphasized by the public health community as essential components for achieving compliance with governmental orders and prevention measures (Blair *et al*. 2017; Freimuth *et al*. 2014; Lazarus *et al*. 2020; Quinn *et al*. 2013; Shore 2003; Siegrist and Zingg 2014). As the experience of the United States, another country with a competitive election in 2020, has shown, the presence of distrust and lack of support for Covid-19 prevention measures shared by significant fractions of the population can be an impediment to an effective pandemic response (Ciancio *et al*. 2020; Soveri *et al*. 2020). Trust is related to cultural values, norms and beliefs that in Malawi are represented by the authority of the village heads (OECD 2013; Quinn *et al*. 2013) and prior research in SSA has shown that local authorities are often seen as more trustful than national institutions and leaders (Kao *et al*. 2021; Vinck *et al*. 2019). This is consistent with our findings that local sources of information (coming from local health personnel, traditional healers, community leaders and/or religious leaders) are more important to implementing social distancing measures than national sources such as newspapers, radio, TV, etc (Supplemental Table S11). Moreover, we show that trust in institutions is positively associated with subjective well-being, self-reported health and mental health outcomes during the pandemic, suggesting that it has a broader impact and plays an important role for individual’s life satisfaction and quality of life during a period of distress and insecurity (Supplemental Table S10).

In interpreting the above results, it is important to acknowledge some limitations. Our analyses do not formally establish causation, which would be very challenging in the context where this study is conducted and the research questions we attempt to answer. Yet, the consistent patterns of findings across multiple outcomes and specifications is suggestive of potential underlying causal pathways. Moreover, all our outcomes, including Covid-19 knowledge and prevention behavior, are necessarily self-reported, and therefore subject to “social desirability” bias. This limitation is inherent as in-person surveys and direct observation were not feasible during the time-period of this study, and indirect measures (e.g., based on cell phone mobility data) are not viable in a rural Malawi context where cell phone ownership continues to be uncommon (albeit growing). Also, notwithstanding the “success” that we highlight in terms of curtailing Covid-19 infections and mortality in Malawi, the social and economic impacts of the pandemic are severe and possibly lasting. For example, a substantial fraction of the MLSFH respondents reduced non-food consumption as a result from Covid-19, and about 61% reported worries about having enough food to eat. For more than half of the interviewed respondents (55%), the economic situation deteriorated compared to the previous year. Worries about access to health care for NCDs and other medical needs such as malaria treatment or HIV/AIDS care were also high.

In summary, behavioral and institutional responses to Covid-19 will continue to remain a central hallmark of the response to the Covid-19 pandemic in SSA LICs for the foreseeable future as vaccines are only slowly reaching rural and poor populations. In this context, our study is important as it documents the role of local and national stakeholders to implement public health measures, monitor and mitigate the social and economic impacts of the Covid-19 pandemic in these rural communities and protect the welfare of their most vulnerable individuals. Specifically, our findings emphasize the importance of local community leadership for shaping behavioral and economic responses to the pandemic in low-income settings such as Malawi: while the government and health care system are entrusted with defining and prioritizing public health policies during Covid-19, local traditional institutions as represented in our case by the traditional authority of the VHs are indispensable for reinforcing their effective implementation. This involvement of affected communities and their governance is essential, not only in the implementation of official public health measures, but also in the application of countermeasures designed to address the social and economic consequences of the pandemic occurring in rapidly changing health environments.

## Data Availability

Data available upon request

https://malawi.pop.upenn.edu/

## Supplemental Materials

**Table S1:** Associations between Covid-19 infections, mortality and seroprevalence with past mortality due to infectious diseases.

|  | Estim. Cumul.<br>Covid-19<br>Infections<br>per 100k | Total<br>Covid-19<br>Deaths<br>per 100k | Estim.<br>Covid-19<br>Sero-<br>prevalence |
| --- | --- | --- | --- |
| **2010 Age-standardized mortality due to** |  |  |  |
| Neglected tropical diseases and malaria | -0.00525***<br>(0.00106) | -0.00604***<br>(0.00110) | -0.00479***<br>(0.00110) |
| HIV and other STIs | 0.00392**<br>(0.00149) | 0.00451***<br>(0.00135) | 0.00432***<br>(0.00131) |
| Respiratory infections and tuberculosis | 0.000231<br>(0.00409) | -0.00307<br>(0.00426) | -0.000939<br>(0.00378) |
| Enteric infections | -0.00460**<br>(0.00219) | -0.00314<br>(0.00333) | -0.00485**<br>(0.00204) |
| Other infectious diseases | 0.00967<br>(0.00718) | 0.00695<br>(0.00826) | 0.00487<br>(0.00774) |
| All infectious diseases combined | 0.00206<br>(0.00241) | 0.00119<br>(0.00248) | 0.00281<br>(0.00227) |

*Note:* Estimates are derived from heightened linear regressions with robust standard errors reported in parentheses; weights are proportional to country's population size (\* $p < 0.1$ , \*\* $p < 0.05$ , \*\*\* $p < 0.01$ ). Cumulative Covid-19 infections, deaths are seroprevalence are provided by IHME and obtained from IHME Covid-19 disease models calibrated to reported Covid-19 data (IHME 2021), and age-standardized death rate from the various infectious diseases and overall is based on the Global Burden of Disease Study (GBD) 2019 (GBD 2019 Diseases and Injuries Collaborators 2020). Each coefficient is obtained from a separate regression model, including the population age structure (proportion of population younger than 15 years old) and the age-standardized overall mortality rate. Similar estimates are obtained if 2000 instead of 2010 predictors are used

**Table S2:**
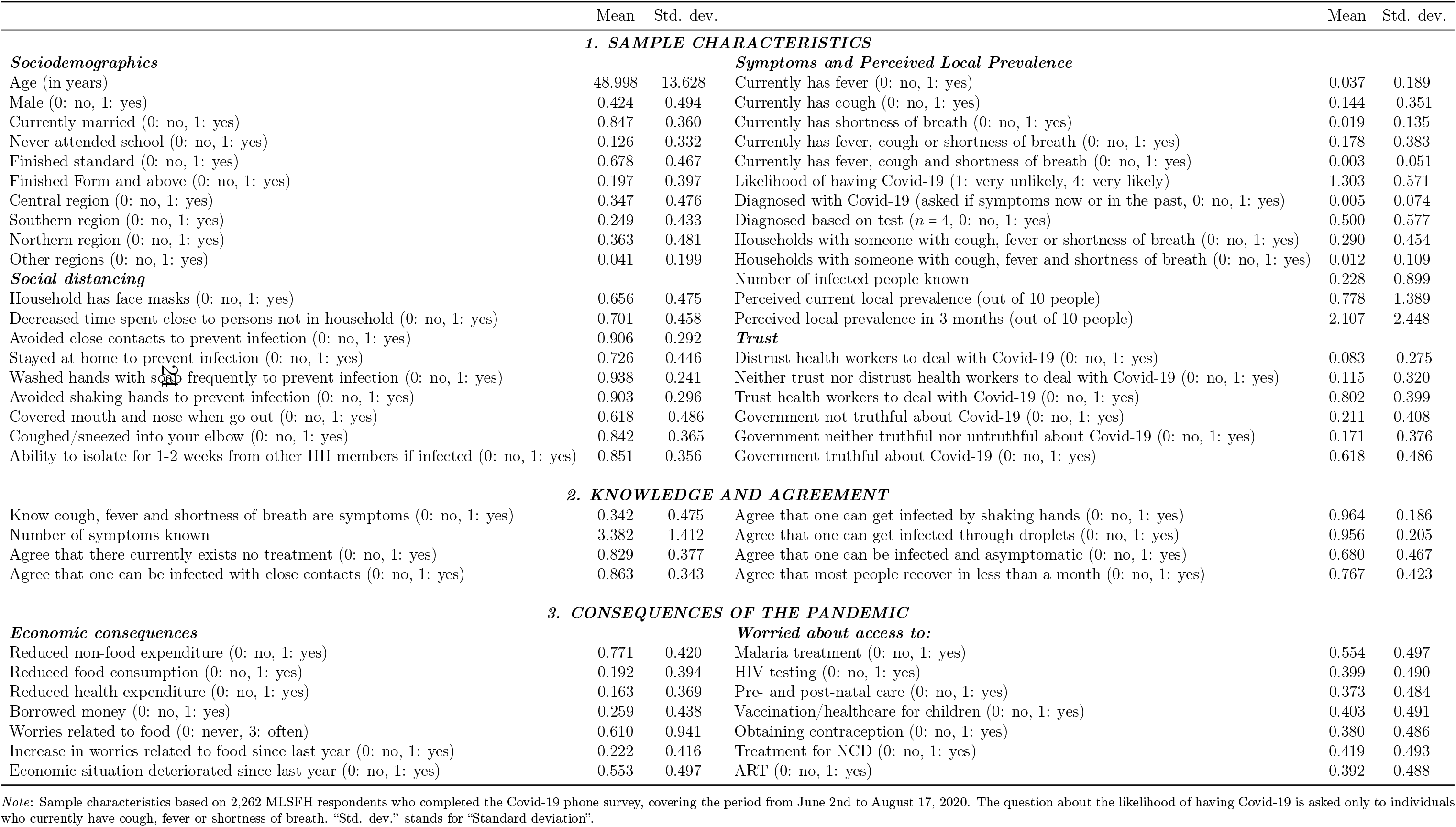
Basic demographics and Covid-19-related characteristics (*N* = 2, 262)

**Table S3:** Associations between village head’s (VH) characteristics and occurrence of negative economic shocks between 2008 and 2010.

|  | Active VH |  |  |  |
| --- | --- | --- | --- | --- |
|  | socially<br>(1) | economically<br>(2) | socially<br>(3) | economically<br>(4) |
| Average number of negative shocks in village | 0.053*<br>(0.031) | -0.009<br>(0.037) |  |  |
| Average number of negative *global* shocks in village |  |  | 0.079**<br>(0.032) | 0.061<br>(0.039) |
| Observations | 2084 | 2084 | 2084 | 2084 |

*Note:* Estimates are derived from linear regressions with robust standard errors reported in parentheses (\* $p < 0.1$ , \*\* $p < 0.05$ , \*\*\* $p < 0.01$ ). All regressions control for sex, age (dichotomous variables for age 19-34, 35-44, 45-54, 55-64, 65-90) and education (dichotomous variables for "never attended school", "finished standard" and "finished form and above"), and include region and time (in days) fixed-effects. We define a VH as being "socially active" if he has instructed respondents to cancel village meetings, keep distance from other people while fetching water, stop public works or stopping recreational activities, such as soccer on the playground. We define a VH as being "economically active" if he has instructed respondents to creating a village fund for emergency purposes or redistribute resources (food, money, medical supplies) to the most vulnerable members of the village community. "Average number of negative shocks in village (2010)" is the average number of economic shocks reported by individuals over the period 2008-2010. "Average number of negative *global* shocks in village (2010)" restrict the economic shocks reported by individuals to have affected other households in the community, and not just respondents'. Analysis is restricted to villages with at least 5 observations (5 respondents).

**Table S4:**
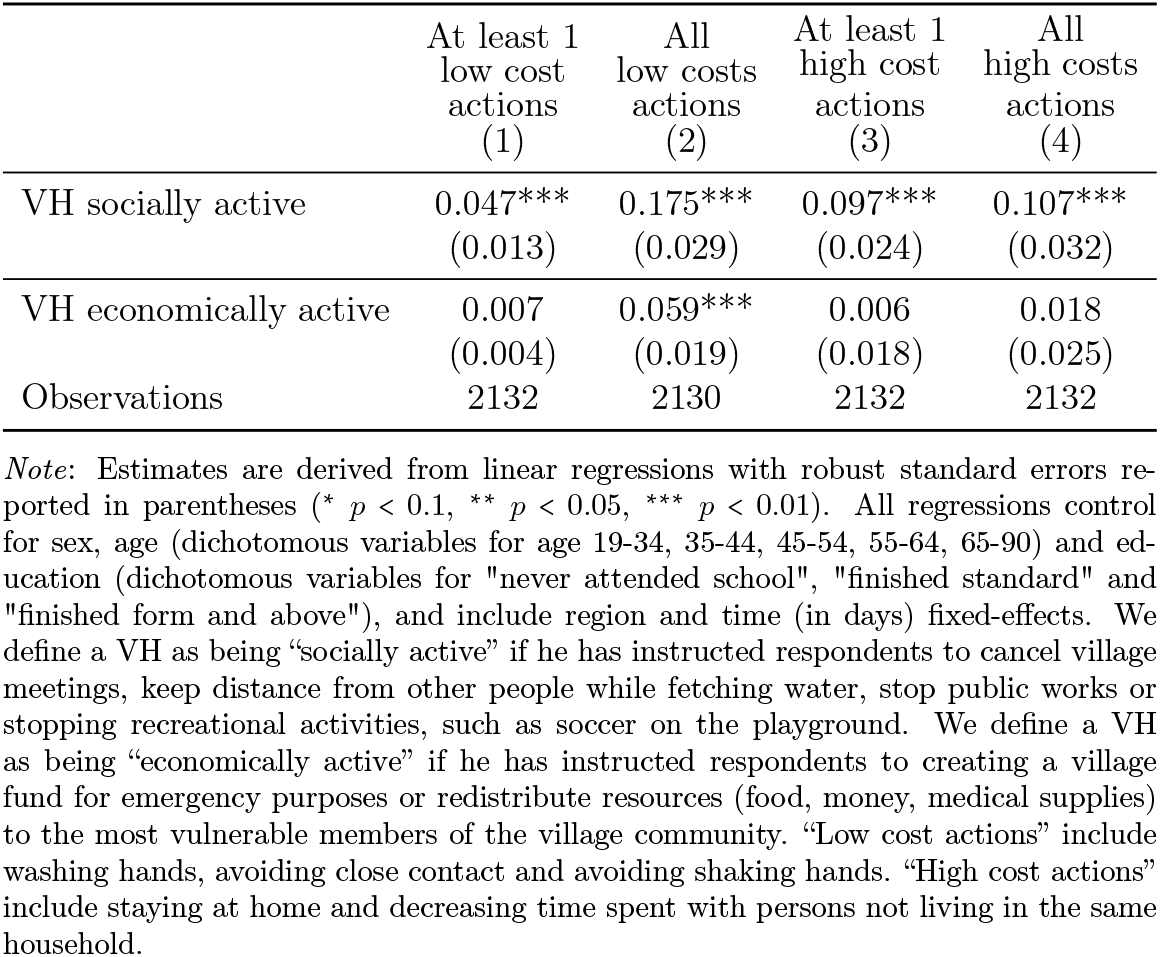
Associations between village head’s (VH) characteristics and actions taken to reduce risks of infection.

|  | At least 1<br>low cost<br>actions<br>(1) | All<br>low costs<br>actions<br>(2) | At least 1<br>high cost<br>actions<br>(3) | All<br>high costs<br>actions<br>(4) |
| --- | --- | --- | --- | --- |
| VH socially active | 0.047***<br>(0.013) | 0.175***<br>(0.029) | 0.097***<br>(0.024) | 0.107***<br>(0.032) |
| VH economically active | 0.007<br>(0.004) | 0.059***<br>(0.019) | 0.006<br>(0.018) | 0.018<br>(0.025) |
| Observations | 2132 | 2130 | 2132 | 2132 |

*Note:* Estimates are derived from linear regressions with robust standard errors reported in parentheses (\* $p < 0.1$ , \*\* $p < 0.05$ , \*\*\* $p < 0.01$ ). All regressions control for sex, age (dichotomous variables for age 19-34, 35-44, 45-54, 55-64, 65-90) and education (dichotomous variables for "never attended school", "finished standard" and "finished form and above"), and include region and time (in days) fixed-effects. We define a VH as being "socially active" if he has instructed respondents to cancel village meetings, keep distance from other people while fetching water, stop public works or stopping recreational activities, such as soccer on the playground. We define a VH as being "economically active" if he has instructed respondents to creating a village fund for emergency purposes or redistribute resources (food, money, medical supplies) to the most vulnerable members of the village community. "Low cost actions" include washing hands, avoiding close contact and avoiding shaking hands. "High cost actions" include staying at home and decreasing time spent with persons not living in the same household.

**Table S5:**
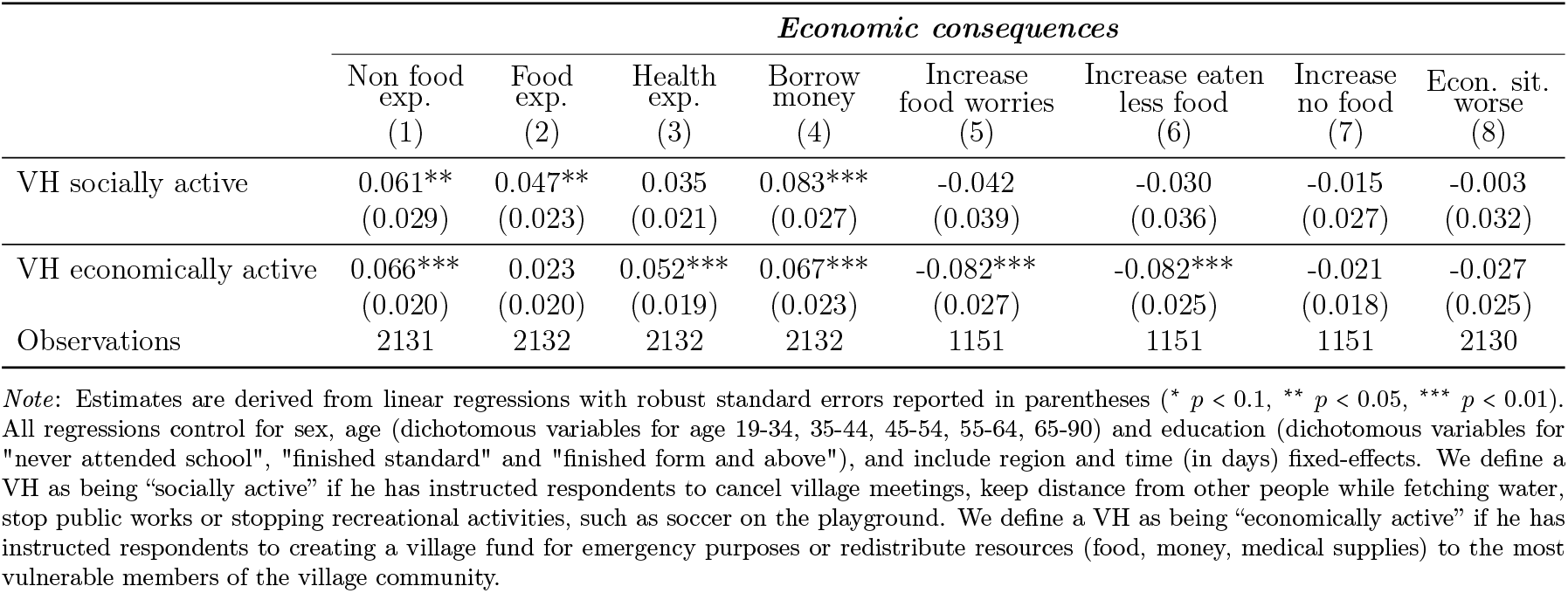
Associations between village head’s (VH) characteristics and economic consequences.

**Table S6:** Associations between village head’s (VH) characteristics and worries about health care access.

|  | *Worries about health care access* |  |  |  |  |  |  |
| --- | --- | --- | --- | --- | --- | --- | --- |
|  | Malaria<br>(1) | HIV testing<br>(2) | Pre/post-natal<br>(3) | Vaccine<br>(4) | Contraception<br>(5) | NCDs<br>(6) | ART<br>(7) |
| VH socially active | 0.069**<br>(0.031) | 0.018<br>(0.031) | -0.007<br>(0.034) | 0.042<br>(0.032) | 0.034<br>(0.033) | 0.020<br>(0.031) | 0.049<br>(0.031) |
| VH economically active | -0.004<br>(0.026) | -0.055**<br>(0.024) | -0.079***<br>(0.026) | -0.084***<br>(0.025) | -0.048*<br>(0.026) | -0.018<br>(0.025) | -0.023<br>(0.025) |
| Observations | 2129 | 2127 | 1875 | 1991 | 1856 | 2096 | 2006 |

*Note:* Estimates are derived from linear regressions with robust standard errors reported in parentheses (\* $p < 0.1$ , \*\* $p < 0.05$ , \*\*\* $p < 0.01$ ). All regressions control for sex, age (dichotomous variables for age 19-34, 35-44, 45-54, 55-64, 65-90) and education (dichotomous variables for "never attended school", "finished standard" and "finished form and above"), an include region and time (in days) fixed-effects. We define a VH as being "socially active" if he has instructed respondents to cancel village meetings, keep distance from other people while fetching water, stop public works or stopping recreational activities, such as soccer on the playground. We define a VH as being "economically active" if he has instructed respondents to creating a village fund for emergency purposes or redistribute resources (food, money, medical supplies) to the most vulnerable members of the village community. "NCDs" stands for non-communicable diseases. "ART" stands for antiretroviral treatment.

**Table S7:**
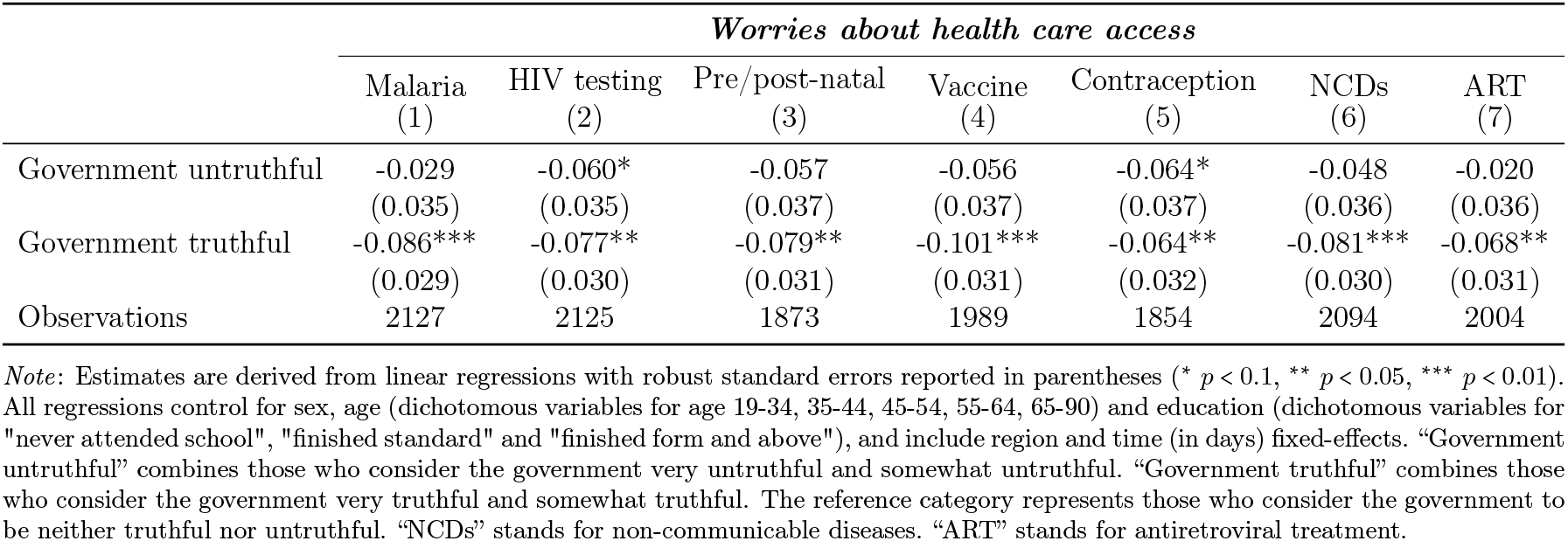
Associations between trust towards institutions and worries about health care access.

**Table S8:** Associations between trust towards institutions (government) and social distancing measures.

|  | *Social distancing* |  |  |  |  |  |  |  |
| --- | --- | --- | --- | --- | --- | --- | --- | --- |
|  | Action score<br>(1) | Face masks<br>(2) | Decreased time<br>(3) | Avoided contact<br>(4) | Stayed at home<br>(5) | Washed hands<br>(6) | Avoided sh. hands<br>(7) | Prayed<br>(8) |
| Government untruthful | -0.217**<br>(0.087) | -0.078**<br>(0.032) | -0.069**<br>(0.034) | -0.035<br>(0.023) | -0.066**<br>(0.031) | -0.046***<br>(0.016) | -0.027<br>(0.022) | -0.106***<br>(0.032) |
| Government truthful | 0.054<br>(0.068) | 0.016<br>(0.027) | 0.049*<br>(0.028) | 0.030*<br>(0.018) | -0.036<br>(0.026) | -0.030**<br>(0.013) | 0.018<br>(0.018) | -0.085***<br>(0.026) |
| Observations | 2125 | 2129 | 2129 | 2129 | 2129 | 2128 | 2128 | 2129 |

*Note:* Estimates are derived from linear regressions with robust standard errors reported in parentheses (\* $p < 0.1$ , \*\* $p < 0.05$ , \*\*\* $p < 0.01$ ). All regressions control for sex, age (dichotomous variables for age 19-34, 35-44, 45-54, 55-64, 65-90) and education (dichotomous variables for "never attended school", "finished standard" and "finished form and above"), and include region and time (in days) fixed-effects. "Government untruthful" combines those who consider the government very untruthful and somewhat untruthful. "Government truthful" combines those who consider the government very truthful and somewhat truthful. The reference category represents those who consider the government to be neither truthful nor untruthful. "Action score" (ranging from 0 to 6) represents the number of appropriate actions *taken* by respondents to reduce the risk of infections such as washed hands, avoided close contacts, stayed at home, covered mouth/nose, avoided shaking hands and coughed in elbow are considered as appropriate actions.

**Table S9:** Associations between trust towards institutions (healthcare workers - HW) and social distancing measures.

|  | *Social distancing* |  |  |  |  |  |  |  |
| --- | --- | --- | --- | --- | --- | --- | --- | --- |
|  | Action score<br>(1) | Face masks<br>(2) | Decrease time<br>(3) | Avoided contact<br>(4) | Stayed at home<br>(5) | Washed hands<br>(6) | Avoided sh. hands<br>(7) | Prayers<br>(8) |
| Distrust in HW | -0.433***<br>(0.122) | -0.073<br>(0.045) | -0.079*<br>(0.046) | -0.066**<br>(0.033) | -0.090**<br>(0.042) | -0.054**<br>(0.027) | -0.110***<br>(0.029) | 0.039<br>(0.045) |
| Trust in HW | -0.068<br>(0.070) | 0.058*<br>(0.031) | -0.027<br>(0.031) | -0.009<br>(0.020) | -0.086***<br>(0.028) | -0.016<br>(0.016) | -0.053***<br>(0.016) | 0.025<br>(0.033) |
| Observations | 2126 | 2130 | 2130 | 2130 | 2130 | 2129 | 2129 | 2130 |

*Note:* Estimates are derived from linear regressions with robust standard errors reported in parentheses (\* $p < 0.1$ , \*\* $p < 0.05$ , \*\*\* $p < 0.01$ ). All regressions control for sex, age (dichotomous variables for age 19-34, 35-44, 45-54, 55-64, 65-90) and education (dichotomous variables for "never attended school", "finished standard" and "finished form and above") and include region and time (in days) fixed-effects. "Distrust in HW" combines those who strongly distrust and those who somewhat distrust HW. "Trust in HW" combines those who strongly trust and those who somewhat trust HW. The reference category represents those who neither trust nor distrust. "Action score" (ranging from 0 to 6) represents the number of appropriate actions *taken* by respondents to reduce the risk of infections such as washed hands, avoided close contacts, stayed at home, covered mouth/nose, avoided shaking hands and coughed in elbow are considered as appropriate actions.

**Table S10:**
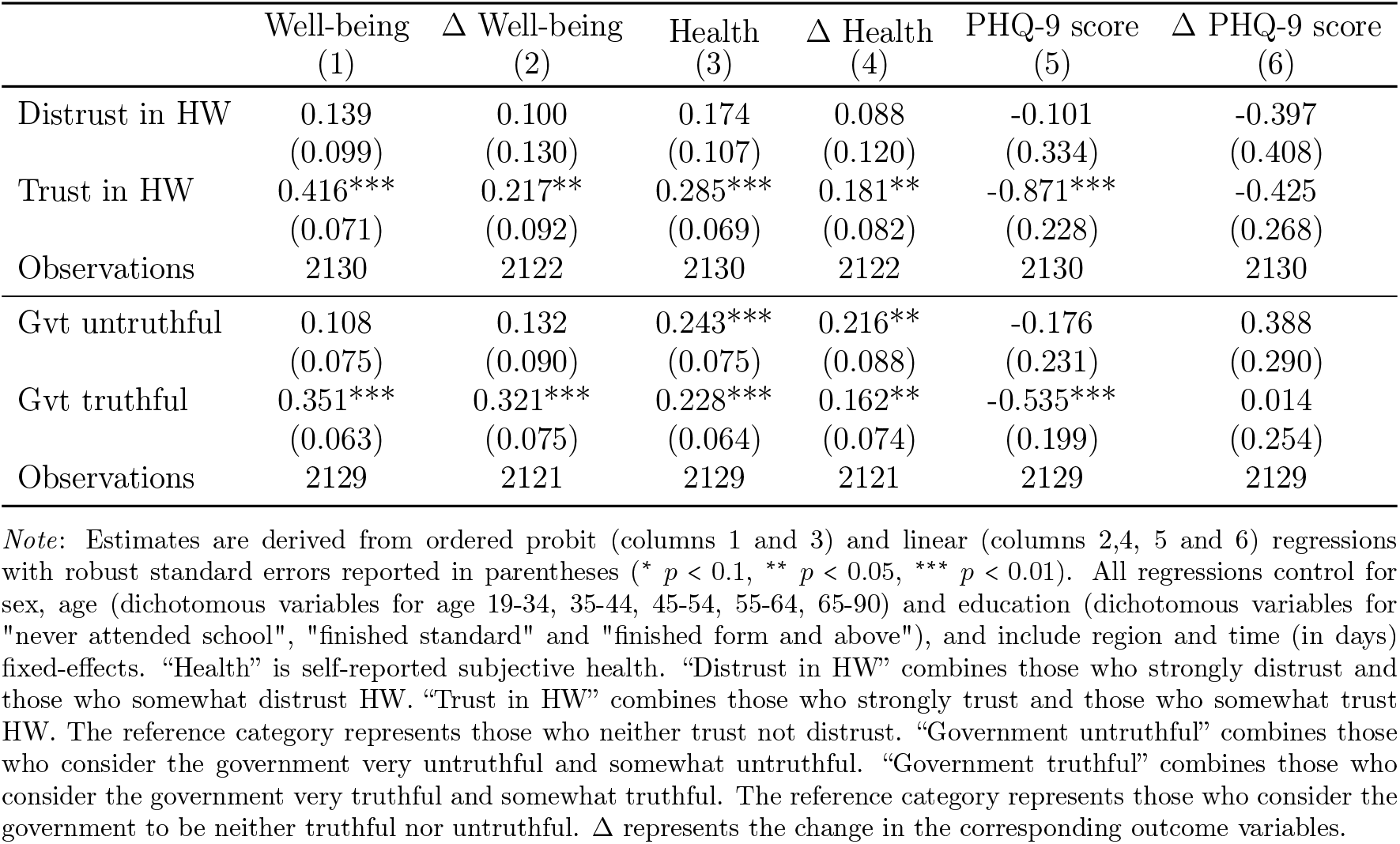
Associations between trust towards institutions and respondent’s wellbeing, self-reported health and mental health (PHQ-9 score)

**Table S11:** Associations between sources of information and social distancing measures.

|  | Wear<br>face masks<br>(1) | HH owns<br>face masks<br>(2) | RR<br>score<br>(3) | Action<br>score<br>(4) | Wear<br>face masks<br>(5) | HH owns<br>face masks<br>(6) | RR<br>score<br>(7) | Action<br>score<br>(8) |
| --- | --- | --- | --- | --- | --- | --- | --- | --- |
| Local source | 0.032<br>(0.025) | 0.029<br>(0.024) | 0.134***<br>(0.049) | 0.235***<br>(0.068) |  |  |  |  |
| National source |  |  |  |  | 0.032<br>(0.024) | 0.040*<br>(0.023) | 0.077<br>(0.049) | 0.167**<br>(0.066) |
| Observations | 2132 | 2133 | 2128 | 2129 | 2132 | 2133 | 2128 | 2129 |

## References

A Madhi, SE Gray, G., Ismail, N., Izu, A., Mendelson, M., Cassim, N., Stevens, W., and Venter, F. (2020). Covid-19 lockdowns in low-and middle-income countries: success against COVID-19 at the price of greater costs. SAMJ: South African Medical Journal, 110(8), 724–726.

Bamgboye, E. L., Omiye, J. A., Afolaranmi, O. J., Davids, M. R., Tannor, E. K., Wadee, S., Niang, A., Were, A., and Naicker, S. (2020). Covid-19 pandemic: Is africa different? Journal of the National Medical Association.

Banda, J., Dube, A., Brumfield, S., Amoah, A., Crampin, A., Reniers, G., and Helleringer, S. (2020). Knowledge and behaviors related to the COVID-19 pandemic in Malawi. medRxiv.

BBC News (2020). Lazarus Chakwera sworn in as Malawi president after historic win. BBC News.

Bearak, M. and Paquette, D. (2020). The coronavirus is ravaging the world. but life looks almost normal in much of Africa. The Washington Post.

Blair, R. A., Morse, B. S., and Tsai, L. L. (2017). Public health and public trust: Survey evidence from the ebola virus disease epidemic in liberia. Social Science & Medicine, 172, 89–97.

BMGF (2020). Covid-19: A global perspective 2020 goalkeepers report. Bill and Melinda Gates Foundation Global Report.

Buonsenso, D., Cinicola, B., Raffaelli, F., Sollena, P., and Iodice, F. (2020). Social consequences of COVID-19 in a low resource setting in Sierra Leone, West Africa. International Journal of Infectious Diseases, 97, 23–26.

Burger, R., Nkonki, L., Rensburg, R., Smith, A., and van Schalkwyk, C. (2020). Examining the unintended health consequences of the COVID-19 pandemic in South Africa.

Chan, M. (2014). Ebola virus disease in West Africa—no early end to the outbreak. New England Journal of Medicine, 371(13), 1183–1185.

Chirwa, G. C., Dulani, B., Sithole, L., Chunga, J. J., Alfonso, W., and Tengatenga, J. (2020). Malawi at the crossroads: Does the fear of contracting covid-19 affect the propensity to vote? The European Journal of Development Research, pages 1–23.

Ciancio, A., Kämpfen, F., Kohler, I. V., Bennett, D., Bruine de Bruin, W., Darling, J., Kapteyn, A., Maurer, J., and Kohler, H.-P. (2020). Know your epidemic, know your response: Early perceptions of COVID-19 and self-reported social distancing in the United States. PLOS ONE. Published September 4, 2020.

Delavande, A. and Kohler, H.-P. (2009). Subjective expectations in the context of HIV/AIDS in Malawi. Demographic Research, 20(31), 817–874.

Egger, D., Miguel, E., Warren, S. S., Shenoy, A., Collins, E., Karlan, D., Parkerson, D., Mobarak, A. M., Fink, G., Udry, C., et al. (2021). Falling living standards during the covid-19 crisis: Quantitative evidence from nine developing countries. Science Advances, 7(6), eabe0997.

Fitzpatrick, A. E., Beg, S. A., Derksen, L. C., Karing, A., Kerwin, J. T., Lucas, A., Reynoso, N. O., and Squires, M. (2021). Health knowledge and nonpharmaceutical interventions during the COVID-19 pandemic in Africa. Technical report, National Bureau of Economic Research.

Freimuth, V. S., Musa, D., Hilyard, K., Quinn, S. C., and Kim, K. (2014). Trust during the early stages of the 2009 h1n1 pandemic. Journal of Health Communication, 19(3), 321–339. PMID: 24117390.

Gates, B. (2020). Responding to Covid-19—a once-in-a-century pandemic? New England Journal of Medicine, 382(18), 1677–1679.

GBD 2019 Diseases and Injuries Collaborators (2020). Global burden of 369 diseases and injuries in 204 countries and territories, 1990–2019: a systematic analysis for the global burden of disease study 2019. Lancet, 396(10258), 1204–1222.

Hargreaves, J., Davey, C., Auerbach, J., Blanchard, J., Bond, V., Bonell, C., Burgess, R., Busza, J., Colbourn, T., Cowan, F., et al. (2020). Three lessons for the COVID-19 response from pandemic HIV. The Lancet HIV, 7(5), e309– e311.

Hopman, J., Allegranzi, B., and Mehtar, S. (2020). Managing COVID-19 in low-and middle-income countries. JAMA, 323(16), 1549–1550.

IHME (2021). COVID-19 mortality, infection, testing, hospital resource use, and social distancing projections. Institute for Health Metrics and Evaluation (IHME), University of Washington, Data Version March 6, 2021.

Kanu, I. A. (2020). Covid-19 and the economy: an African perspective. Journal of African Studies and Sustainable Development, 3(2).

Kao, K., Lust, E., Dulani, B., Ferree, K. E., Harris, A. S., and Metheney, E. (2021). The ABCs of Covid-19 prevention in Malawi: Authority, benefits, and costs of compliance. World development, 137, 105167.

Kohler, H.-P., Watkins, S. C., Behrman, J. R., Anglewicz, P., Kohler, I. V., Thornton, R. L., Mkandawire, J., Honde, H., Hawara, A., Chilima, B., Bandawe, C., and Mwapasa, V. (2015). Cohort profile: The Malawi Longitudinal Study of Families and Health (MLSFH). International Journal of Epidemiology, 44(2), 394–404.

Lazarus, J. V., Binagwaho, A., El-Mohandes, A. A. E., Fielding, J. E., Larson, H. J., Plaséncia, A., Andriukaitis, V., and Ratzan, S. C. (2020). Keeping governments accountable: the COVID-19 assessment scorecard (COVIDSCORE). Nature Medicine, 26(7), 1005–1008.

Maeda, J. M. and Nkengasong, J. N. (2021). The puzzle of the COVID-19 pandemic in Africa. Science, 371(6524), 27–28.

Mbow, M., Lell, B., Jochems, S. P., Cisse, B., Mboup, S., Dewals, B. G., Jaye, A., Dieye, A., and Yazdanbakhsh, M. (2020). Covid-19 in africa: Dampening the storm? Science, 369(6504), 624–626.

Mukherjee, S. (2021). The Covid conundrum: Why does the pandemic seem to be hitting some countries harder than others? The New Yorker. Published in the print edition of the March 1, 2021.

Musa, H. H., Musa, T. H., Musa, I. H., Musa, I. H., Ranciaro, A., and Campbell, M. C. (2021). Addressing Africaâs pandemic puzzle: Perspectives on COVID-19 transmission and mortality in sub-Saharan Africa. International Journal of Infectious Diseases, 102, 483–488.

Nuwagira, E. and Muzoora, C. (2020). Is sub-Saharan Africa prepared for COVID-19? Tropical Medicine and Health, 48(1), 1–3.

Oecd, O. (2013). Trust in government, policy effectiveness and the governance agenda. Government at a Glance, 2013.

Paintsil, E. et al. (2020). Covid-19 threatens health systems in sub-saharan africa: the eye of the crocodile. The journal of clinical investigation, 130(6).

Public Health Institute of Malawi (2020). COVID-19 daily situation report. Ministry of Health - Malawi.

Quinn, S. C., Parmer, J., Freimuth, V. S., Hilyard, K. M., Musa, D., and Kim, K. H. (2013). Exploring communication, trust in government, and vaccination intention later in the 2009 h1n1 pandemic: results of a national survey. Biosecurity and bioterrorism: biodefense strategy, practice, and science, 11(2), 96–106.

Roberton, T., Carter, E. D., Chou, V. B., Stegmuller, A. R., Jackson, B. D., Tam, Y., Sawadogo-Lewis, T., and Walker, N. (2020). Early estimates of the indirect effects of the covid-19 pandemic on maternal and child mortality in low-income and middle-income countries: a modelling study. The Lancet Global Health, 8(7), e901 – e908.

Rosenthal, P. J., Breman, J. G., Djimde, A. A., John, C. C., Kamya, M. R., Leke, R. G., Moeti, M. R., Nkengasong, J., and Bausch, D. G. (2020). Covid-19: shining the light on Africa. The American Journal of Tropical Medicine and Hygiene, 102(6), 1145.

Shore, D. (2003). Communicating in times of uncertainty: the need for trust. Jounral of Health Communications, 8(Suppl. 1), 13–14.

Shuchman, M. (2020). Low-and middle-income countries face up to COVID-19. Nat. med.

Siegrist, M. and Zingg, A. (2014). The role of public trust during pandemics: Implications for crisis communication. European Psychologist, 19(1), 23.

Soveri, A., Karlsson, L. C., Antfolk, J., Lindfelt, M., and Lewandowsky, S. (2020). Unwillingness to engage in behaviors that protect against COVID-19: Conspiracy, trust, reactance, and endorsement of complementary and alternative medicine.

The Republic of Malawi (2020). National covid-19 preparedness and response plan.

UNDP (2000). The next frontier - Human development and the Anthropocene.

Van Zandvoort, K., Jarvis, C. I., Pearson, C. A., Davies, N. G., Ratnayake, R., Russell, T. W., Kucharski, A. J., Jit, M., Flasche, S., Eggo, R. M., et al. (2020). Response strategies for COVID-19 epidemics in African settings: a mathematical modelling study. BMC medicine, 18(1), 1–19.

Vinck, P., Pham, P. N., Bindu, K. K., Bedford, J., and Nilles, E. J. (2019). Institutional trust and misinformation in the response to the 2018–19 Ebola outbreak in North Kivu, DR Congo: a population-based survey. The Lancet Infectious Diseases, 19(5), 529–536.

Walker, P. G., Whittaker, C., Watson, O. J., Baguelin, M., Winskill, P., Hamlet, A., Djafaara, B. A., Cucunubá, Z., Mesa, D. O., Green, W., et al. (2020). The impact of covid-19 and strategies for mitigation and suppression in low-and middle-income countries. Science, 369(6502), 413–422.

WHO (2020). Coronavirus disease 2019 (COVID-19): Situation report, 198. Worldometers (2020). Covid-19 coronavirus pandemic. Accessed: 2020-09-24.

